# Perceived Challenges of COVID-19 Infection Prevention and Control Preparedness: A Multinational Survey

**DOI:** 10.1101/2020.06.17.20133348

**Authors:** Ermira Tartari, Joost Hopman, Benedetta Allegranzi, Bin Gao, Andreas Widmer, Vincent Chi-Chung Cheng, Shuk Ching Wong, Kalisvar Marimuthu, Folasade Ogunsola, Andreas Voss, on behalf of the International Society of Antimicrobial Chemotherapy Infection and Prevention Control (ISAC-IPC) Working Group

## Abstract

**Objectives:** Implementation of effective infection prevention and control (IPC) measures is needed to support global capacity building to limit transmission of coronavirus disease 2019 (COVID-19) and mitigate its impact on health systems. We assessed the perceptions of healthcare workers on the current global IPC preparedness measures for COVID-19.

**Methods:** A cross-sectional survey using an electronic survey was circulated between February 26, 2020, and March 20, 2020, to IPC professionals during COVID-19 pandemic. The survey addressed the presence of COVID-19 guidelines as well as specific IPC preparedness activities in response to the outbreak.

**Findings:** In total, 339 IPC professionals spanning 63 countries in all 6 World Health Organization (WHO) regions, mostly from tertiary care centres participated. Of all participants, 66·6% were aware of the existence of national guidelines to prevent COVID-19. A shortage of PPE supplies was reported by 48% (ranging from 64·2% in low-income countries to 27·4% in high-income countries); 41·5% of respondents considered that the media had an impact on guideline development and 63·6% believed that guidelines were based on maximum security rather than on evidence-based analyses. 58·5% and 72·7% of participants believed that healthcare facilities and community settings respectively were not sufficiently prepared.

**Conclusion:** Results revealed lack of guidelines and concerns over insufficient PPE supply in both high- and low-income countries. Our findings should alert national health authorities to ramp up the implementation of IPC measures and focus on long-term preparedness and readiness for future pandemics, likely requiring government funds rather than reliance on healthcare institutions.

## Introduction

The emergence of the unprecedented novel coronavirus disease 2019 (COVID-19) outbreak in China[1,2] and its rapid spread worldwide is of major global concern to public health and national economies.[3] On January 30, 2020, the World Health Organization (WHO) declared COVID-19 as a public health emergency of international concern under the International Health Regulations (2005)[4] and on March 11, 2020, COVID-19 was declared a global pandemic.[5] As of 19 April, 2020, over 7 390 702 cases and 417 731 deaths have been reported across the globe.[3]

Acknowledgment of the major threat represented by this new virus prompted WHO to convene a global research roadmap forum on 11/12 February 2020 with leading health experts to assess the evidence about COVID-19 disease, identify gaps and prioritize research needed in a number of critical areas, to make rapid progress in the fight against the virus.[6] Infection prevention and control (IPC) is one of the critical areas identified as a priority for rapid research action. Since the beginning of the outbreak, WHO’s leadership recommended all countries to rapidly develop national preparedness capacities for the detection, investigation and management of suspected or confirmed COVID-19 cases and to implement the WHO infection prevention and control (IPC) core components in order to respond effectively.[7]

Previous experience of outbreaks due to the severe acute respiratory syndrome coronavirus (SARS-CoV), Ebola, and the Middle East respiratory syndrome coronavirus (MERS-CoV) including the ongoing large-scale outbreak of COVID-19, has shown a high incidence of transmissibility of health care-associated infections and outbreaks affecting healthcare workers (HCWs) who are at the forefront of these crises, illustrating the importance of being prepared.[8–11] Up to 24 February 2020 (day 56), the National Health Commission of the People’s Republic of China reported that 3387 HCWs were infected with COVID-19, resulting in 22 (0·6%) deaths.[12] In Italy, according to the estimates of the National Public Health Institute, as of April 15, out of a total of 155 467 positive cases, 16 650 were HCWs,[13] with the numbers increasing over time.

## Methods

### Objectives

The main objectives of the study were to gain a rapid insight into the preparedness of healthcare facilities and investigate current global practices and perceptions among IPC professionals concerning the prevention and control of COVID-19 so as to identify opportunities for improving practices globally.

### Setting and participants

From February 26 through March 20, 2020, the International Society of Antimicrobial Chemotherapy (ISAC) Working Group for IPC in partnership with Infection Control Africa Network (ICAN) launched a cross-sectional, self-administered web-based survey of IPC specialists working in healthcare facilities to explore IPC measures for COVID-19 preparedness. ISAC Working Group for IPC has affiliated members and IPC representatives from 57 countries worldwide. The IPC Working Group has been initiated to support international research projects and for standardization of guidelines and IPC measures across countries worldwide. To achieve coverage across low- and middle-income countries, agreement was reached to form a working party with ICAN. An online link to the electronic questionnaire was distributed through ISAC and ICAN members and networks and also via social media platforms inviting IPC experts from each continent and country to participate. The link to the web-based survey was made accessible on the societies’ websites. IPC specialists including microbiologists, infectious diseases physicians, nurses, antimicrobial pharmacists and other specialists working in healthcare facilities preparing for the detection, investigation and management of confirmed and suspected COVID-19 patients were invited to participate. Participants were enrolled through a non-random, convenience sampling method. Due to the non-probabilistic sampling nature, it was not possible to determine the statistical representatives of the sample.

The participants enrolled by a web-survey where information about the study’s purpose was provided. Participants were informed that the purpose of the survey was intended to help inform the current progress of countries with IPC measures and preparedness plans for preventing COVID-19. There were no exclusion criteria.

Participation in this study was voluntary and anonymous. Participants completed a web-based questionnaire tool, generated using Survey Monkey platform (SurveyMonkey®, San Mateo, CA, USA). Pre-testing of the platform was performed with 20 users from all six regions to evaluate usability and detect technical failures. The study was exempted by the Radboud University Medical Center (The Netherlands) as it did not fall within the remit of the Medical Research Involving Human Subjects Act (NL2020-6262). The study follows the Strengthening the Reporting of Observational Studies in Epidemiology (STROBE) reporting guideline (Supplementary file 1).

### Survey design, administration and analysis

The survey instrument was a self-designed questionnaire, compiled by IPC scientists and experts participating in the WHO COVID-19 global research and innovation roadmap forum and based on the priorities and thematic areas identified at the forum.[6] After assessing content validity, the instrument was pilot tested among 10 IPC opinion leaders in six countries, in three WHO regions (Europe, Africa and South-East Asia) to evaluate its acceptance and reproducibility. The questionnaire was created in English to gain insight into the preparedness of healthcare facilities. Two study investigators translated the English version of the instrument to Chinese. To confirm that the Chinese translation was reliable, the instrument was back translated to English. The final instrument (Supplementary file 2) was available in English and Chinese. The questionnaire included 47 items in total covering six dimensions addressing: 1) geographic location, demographics and healthcare facility characteristics; 2) the presence of national/regional/local COVID-19 guidelines; 3) personal protective equipment (PPE) type and supplies in healthcare facilities; 4) the role of environmental contamination; 5) preparedness for COVID-19 activities; and 6) the influence of media on preparedness plans. The instrument included dichotomous (yes/no) closed-ended questions and for one section participants were asked to provide their degree of agreement with each item (Agree and Disagree). All responses to questions were allocated a numerical score on the survey platform. Submission of responses via the online platform (SurveyMonkey®, San Mateo, CA, USA) indicated agreement to participate. It was decided to use all entries that contained information about COVID-19 IPC measures and activities, even if there were missing responses for some of the items. Entries with no demographic information were excluded.

### Statistical Analysis

Descriptive statistics were used to analyse the survey data. For descriptive purposes, medians with IQR, cumulative frequencies or 95% confidence intervals (CI) were calculated where appropriate. Means and ranges were aggregated at UN regional level (Africa, the Americas, Eastern Mediterranean, Europe, Southeast Asia, Western Pacific) and country income group according to the World Bank classification (high-income, upper-middle-income, lower-middle-income and low-income).[14] A p-value of less than 0.05 was considered significant. Analyses were performed using R version 3.5.1 (R Core Team. R: A language and environment for statistical computing. R Foundation for Statistical Computing, Vienna, Austria; 2017; https://www.R-project.org/).

## Results

### Characteristics of Survey Respondents

A total of 349 responses were received; 10 were excluded as no demographic information was provided. A sample of 349 participants selected from the two organizations which include approximately 800 IPC specialists guarantees a maximum margin of error of 3.94% assuming a 95% confidence level. The 339 eligible responses were coming from 63 countries across the 6 regions: Africa, 113; Europe, 92; Southeast Asia, 72; the Americas, 33; Eastern Mediterranean, 15; Western Pacific, 14). They represented 113 responses from high-income countries (HICs), 99 from upper-middle-income countries (UMICs), 71 from lower-middle-income countries (LMICs) and 56 from low-income countries (LICs). Response rate by profession included 190 IPC physicians (56·0%); 113 IPC nurses (33·3%) and 36 other professionals, including pharmacists and public health specialists. Healthcare facilities represented in the survey were mostly tertiary care centres (46%) (Table 1).

**Table 1.**
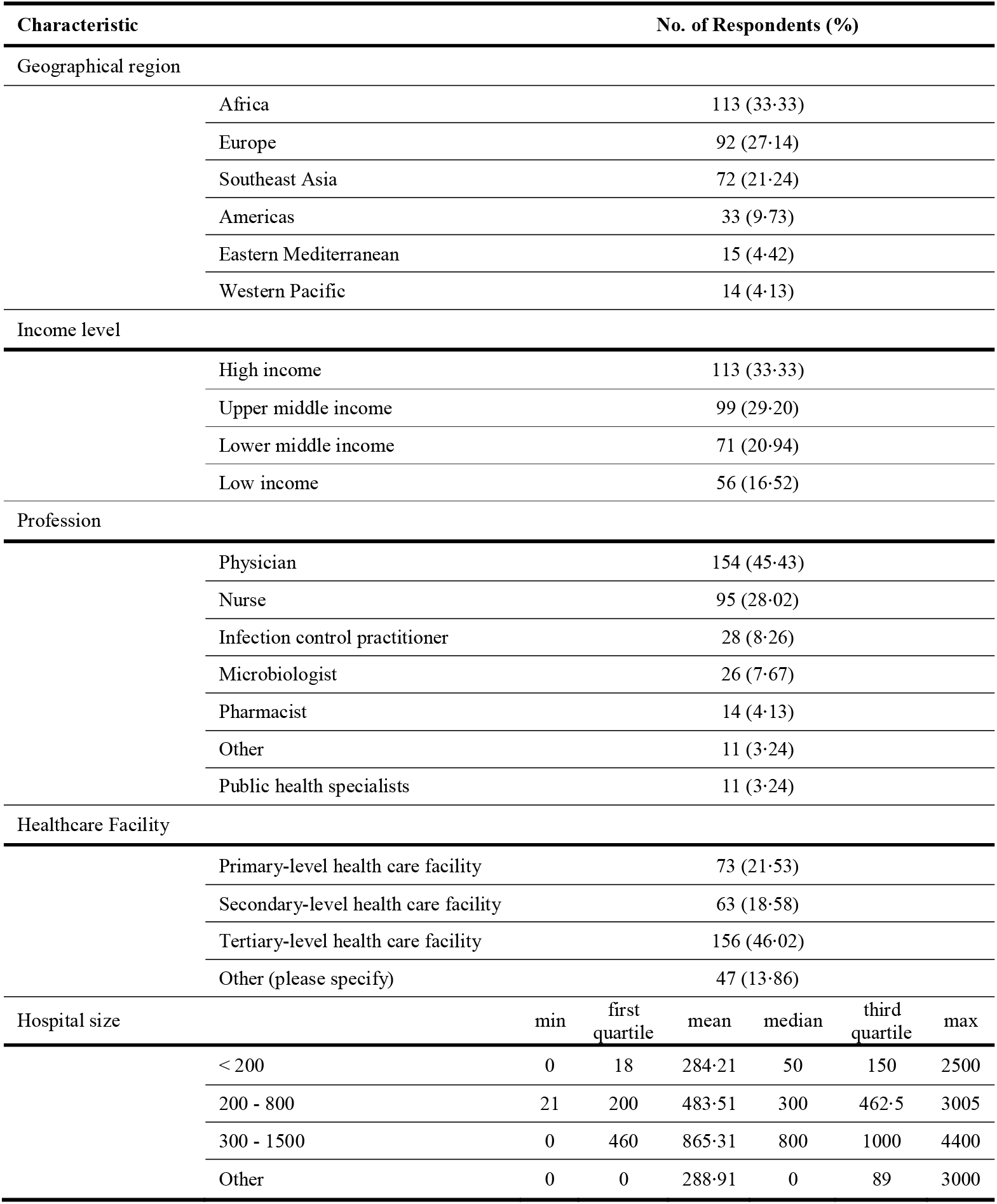
**Characteristics of Survey Respondents**

### COVID-19 Guidelines

Respondents reported that either national (226/339; 66·6%; 95% CI, 61·6-71·6), local or regional (182/339; 53·6%; 95% CI, 48·3-59) guidelines on the prevention and management of COVID-19 were available, but with a significant variation according to region/income level, e.g. HICs (65·5%; 95% CI, 48·2-82·8) were more likely to have national guidelines available than LICs (7·6%; 95% CI, 0-22·1) (*P* <·01) (figure 1; Table 2). When guidelines were not available, respondents mainly adopted international guidelines (72/106; 67·9%). Adopted guidelines were based on those from WHO (42·4%), the European Centre for Disease Prevention and Control (ECDC) 4·7%, Public Health England (PHE) 4·7%, the US Centers for Disease Control and Prevention (CDC) (3·7%) and a combination of WHO, ECDC, CDC and/or PHE (12·2%). The remaining respondents did not know. Almost all respondents (259/269; 96·2%; 95% CI, 94-98·5) affirmed that guidelines included hand hygiene as an important measure (Fig.1, Table 2).

**Table 2.** **Geographical comparison of healthcare facilities and IPC preparedness for patients with COVID-19, results from survey of representatives from 339 healthcare facilities in 63 countries worldwide, February-March**

|  | No. (%) of Respondents | Comparison between regions |  |  |  |  |  | P-value |
| --- | --- | --- | --- | --- | --- | --- | --- | --- |
|  |  | Africa | Americas | East Mediterranean | Europe | South-East Asia | Western Pacific |  |
|  |  | N (%) | N (%) | N (%) | N (%) | N (%) | N (%) |  |
| **COVID-19 Guidelines** |  |  |  |  |  |  |  |  |
| National guidelines | 226 (66·67) | 53 (46·9) | 22 (66·67) | 13 (86·67) | 65 (70·65) | 60 (83·33) | 13 (92·86) | <0·001 |
| Local guidelines | 182 (53·69) | 40 (35·4) | 21 (63·64) | 8 (53·33) | 42 (45·65) | 62 (86·11) | 9 (64·29) | <0·001 |
| Guidelines recommend use of PPE |  |  |  |  |  |  |  |  |
| Guidelines address PPE | 214 (63·13) | 60 (53·1) | 20 (60·61) | 9 (60) | 51 (55·43) | 63 (87·5) | 11 (78·57) | <0·001 |
| Facemask | 272 (80·24) | 87 (76·99) | 25 (75·76) | 11 (73·33) | 70 (76·09) | 65 (90·28) | 14 (100) | 0·06 |
| Gown | 251 (74·04) | 81 (71·68) | 22 (66·67) | 9 (60) | 65 (70·65) | 60 (83·33) | 14 (100) | 0·04 |
| Cap | 182 (64·31) | 66 (68·75) | 8 (29·63) | 5 (50) | 42 (60) | 57 (86·36) | 4 (28·57) | <0·001 |
| Eye protection | 245 (72·27) | 75 (66·37) | 22 (66·67) | 10 (66·67) | 65 (70·65) | 60 (83·33) | 13 (92·86) | 0·07 |
| Gloves | 266 (97·08) | 90 (96·77) | 26 (96·3) | 10 (100) | 65 (95·59) | 62 (98·41) | 13 (100) | 0·004 |
| Preparedness effort |  |  |  |  |  |  |  |  |
| Hand hygiene | 259 (96·28) | 86 (93·48) | 24 (92·31) | 10 (100) | 63 (98·44) | 65 (98·48) | 11 (100) | 0·34 |
| Training HCWs | 235 (86·72) | 67 (72·83) | 24 (85·71) | 10 (100) | 59 (92·19) | 64 (96·97) | 11 (100) | <0·001 |
| PPE in community | 144 (53·33) | 51 (55·43) | 8 (29·63) | 1 (10) | 27 (42·19) | 51 (77·27) | 6 (54·55) | <0·001 |
| PPE in outpatient setting | 243 (91·35) | 80 (86·96) | 21 (84) | 10 (100) | 56 (90·32) | 65 (98·48) | 11 (100) | 0·07 |
| Environmental decontamination |  |  |  |  |  |  |  |  |
| Use of Hypochlorite | 199 (73·7) | 74 (80·43) | 13 (48·15) | 5 (50) | 40 (62·5) | 61 (92·42) | 6 (54·55) | <0·001 |
| Automated disinfection system | 100 (37·04) | 12 (13·04) | 6 (22·22) | 4 (40) | 25 (39·06) | 52 (78·79) | 1 (9·09) | <0·001 |

**Figure 1:**
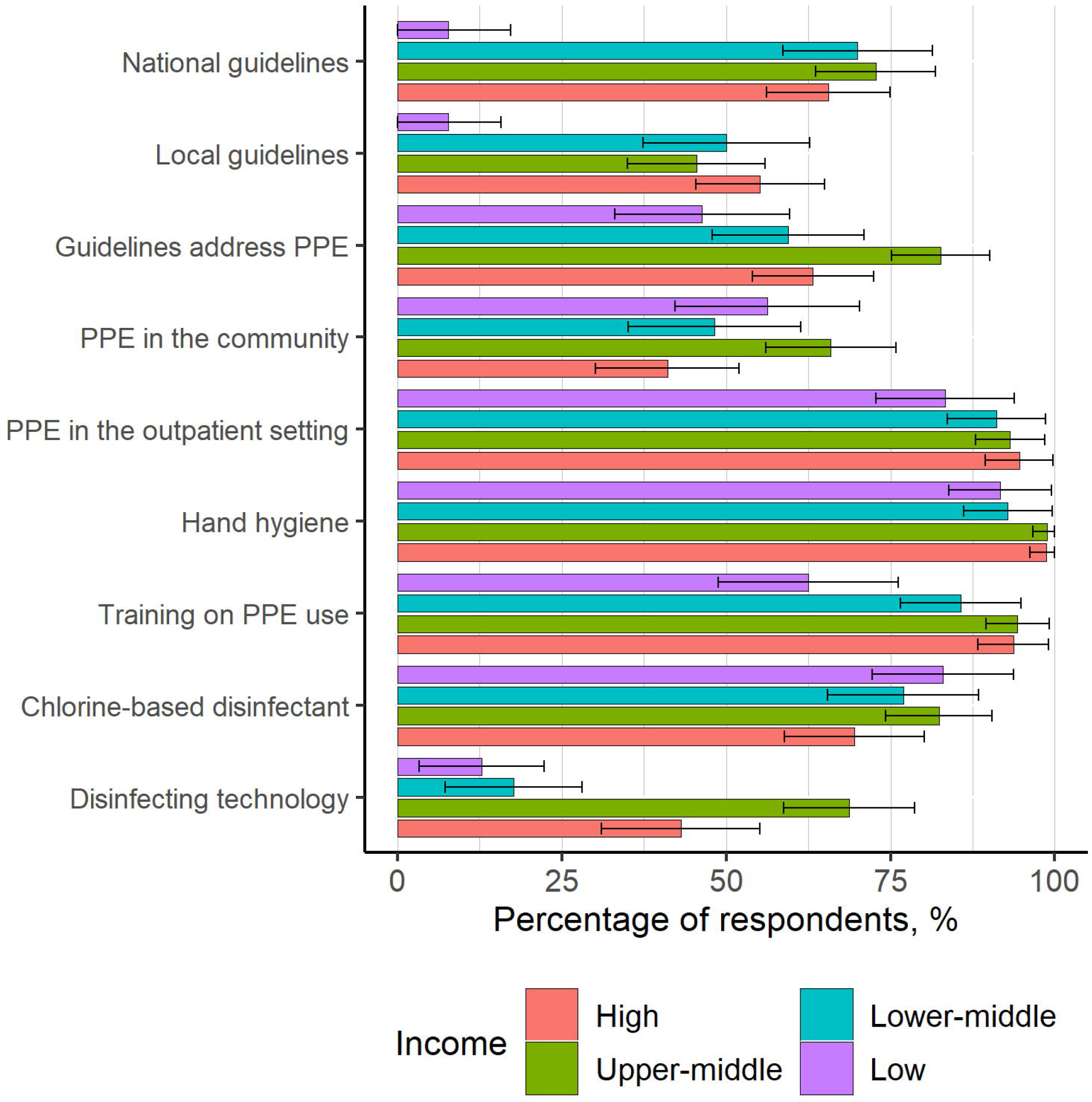
Presence of COVID-19 Guidelines and the Most Critical Infection Prevention and Control Measures Recommended as Identified Among Survey Respondents, Stratified by Country Income Level. HIC, high-income country; UMIC, upper-middle income country; LMIC, lower-middle income country; LIC, low-income country; PPE, personal protective equipment.

### Personal Protective Equipment

More than half (214/339; 63·1%; 95% CI, 57·9-68·2) of respondents reported that guidelines for healthcare facilities addressed PPE use, with a significant variation by region/income level. (Fig. 1, Table 2).

#### Facemasks

Participants reported that national or local COVID-19 guidelines recommended mainly the use of N95/FFP2 respirators (120/267; 44·9%), followed by surgical masks (77/267; 28·8%) or a combination of the two, respectively (39/267; 14·6%), and powered air-purifying respirators (PAPR) (21/267; 7·9%). Variations existed within the 6 regions (Table 3). In Africa and Western Pacific regions, N95/FFP2 respirators and surgical masks (36/85; 42·3% and 6/14; 42·8%, respectively) were mainly recommended. In Europe, the Americas and Eastern Mediterranean, N95/FFP2 respirators were recommended according to more than half of respondents (55·8%, 52% and 70%, respectively). The use of PAPR was recommended in Africa (8/85; 9·4%) and Europe (12/68; 17·6%) *P* = ·15.

**Table 3.** **Protective Equipment (PPE) included in national or local COVID-19 guidelines**

|  | No. (%) of Respondents | Comparison between regions |  |  |  |  |  | P-value |
| --- | --- | --- | --- | --- | --- | --- | --- | --- |
|  |  | Africa | Americas | East Mediterranean | Europe | South-East Asia | Western Pacific |  |
|  | N (%) | N (%) | N (%) | N (%) | N (%) | N (%) | N (%) |  |
| **Face mask** | N = 267 |  |  |  |  |  |  |  |
| FFP1 | 6 (2.25) | 2 (2.35) | 0 (0) | 0 (0) | 3 (4.41) | 1 (1.54) | 0 (0) | 0.16 |
| N95/FFP2 | 120 (44.94) | 36 (42.35) | 13 (52) | 7 (70) | 38 (55.88) | 20 (30.77) | 6 (42.86) | < 0.001 |
| Respirators | 21 (7.87) | 8 (9.41) | 1 (4) | 0 (0) | 12 (17.65) | 0 (0) | 0 (0) | < 0.001 |
| Surgical mask & N95/FFP2 | 39 (14.61) | 1 (1.18) | 1 (4) | 0 (0) | 2 (2.94) | 33 (50.77) | 2 (14.29) | < 0.001 |
| Surgical mask | 77 (28.84) | 36 (42.35) | 10 (40) | 3 (30) | 11 (16.18) | 11 (16.92) | 6 (42.86) | < 0.001 |
| Other | 4 (1.5) | 2 (2.35) | 0 (0) | 0 (0) | 2 (2.94) | 0 (0) | 0 (0) | < 0.001 |
| **Gown type:** | N = 242 |  |  |  |  |  |  |  |
| Short-sleeved plastic | 27 (11.16) | 4 (5.41) | 2 (9.09) | 0 (0) | 1 (1.59) | 20 (33.33) | 0 (0) | < 0.001 |
| Long sleeve water repellant | 170 (70.25) | 52 (70.27) | 20 (90.91) | 8 (88.89) | 55 (87.3) | 23 (38.33) | 12 (85.71) | < 0.001 |
| Coverall | 29 (11.98) | 18 (24.32) | 0 (0) | 1 (11.11) | 7 (11.11) | 1 (1.67) | 2 (14.29) | < 0.001 |
| Long-sleeved-water resistant & Short-sleeved plastic | 16 (6.61) | 0 (0) | 0 (0) | 0 (0) | 0 (0) | 16 (26.67) | 0 (0) | < 0.001 |
| **Cap type:** | N = 181 |  |  |  |  |  |  |  |
| Cap covering head and neck | 79 (43.65) | 44 (67.69) | 4 (50) | 2 (40) | 19 (45.24) | 6 (10.53) | 4 (100) | < 0.001 |
| Cap covering the head only | 102 (56.35) | 21 (32.31) | 4 (50) | 3 (60) | 23 (54.76) | 51 (89.47) | 0 (0) | < 0.001 |
| **Eye protection:** | N = 241 |  |  |  |  |  |  |  |
| "Ski"-goggles | 68 (28.22) | 17 (23.29) | 5 (22.73) | 1 (10) | 15 (23.44) | 29 (48.33) | 1 (8.33) | < 0.001 |
| Face shield | 135 (56.02) | 47 (64.38) | 8 (36.36) | 8 (80) | 37 (57.81) | 28 (46.67) | 7 (58.33) | < 0.001 |
| Other | 38 (15.77) | 9 (12.33) | 9 (40.91) | 1 (10) | 12 (18.75) | 3 (5) | 4 (33.33) | < 0.001 |
| **Gloves:** | N = 274 |  |  |  |  |  |  |  |
| No gloves | 8 (2.92) | 3 (3.23) | 1 (3.7) | 0 (0) | 3 (4.41) | 1 (1.59) | 0 (0) | 0.22 |
| Double gloving | 48 (17.52) | 28 (30.11) | 1 (3.7) | 1 (10) | 14 (20.59) | 2 (3.17) | 2 (15.38) | < 0.001 |
| Single pair disposable | 218 (79.56) | 62 (66.67) | 25 (92.59) | 9 (90) | 51 (75) | 60 (95.24) | 11 (84.62) | < 0.001 |
| **Shoe cover:** | N = 275 |  |  |  |  |  |  |  |
| Shoe and lower leg cover | 50 (18.18) | 30 (32.26) | 0 (0) | 0 (0) | 11 (16.18) | 5 (7.81) | 4 (30.77) | < 0.001 |
| Shoe cover | 99 (36) | 30 (32.26) | 6 (22.22) | 2 (20) | 16 (23.53) | 45 (70.31) | 0 (0) | < 0.001 |
| No shoe cover | 126 (45.82) | 33 (35.49) | 21 (77.77) | 8 (80) | 41 (60.29) | 14 (21.87) | 9 (69.23) | < 0.001 |

PPE: Personal protective equipment; PAPR: powered air-purifying respirators.

#### Gown

Most respondents reported the use of long-sleeve water-repellent or resistant gowns (170/242; 70%). By contrast, Southeast Asia reported a variety of recommended gowns to be used, i.e. long-sleeve water-resistant (38·3%), short-sleeved plastic gown and a combination of both long-sleeve water-resistant and short-sleeved plastic gown (26·6%) concurrently (Table 3).

#### Head protection

More than half of respondents (56·3%) reported the use of a cap that covers the head only, while 43·6% used a cap that covers both head and neck (Table 3). There was a significant regional variation in their recommendations for the use of had protection (*P* <·01) (Table 3).

#### Eye protection

Overall, guidelines recommended the use of eye protection (72·2%; 95% CI, 68-77) (Table 2). More than half (56%) of respondents reported the use of a disposable face shield while 28·2% recommended the use of goggles in the guidelines with significant regional differences (*P* <·01) (Table 3).

#### Gloves

The highest recommendation was reported for single-use disposable gloves (79·5%). By contrast, double gloving was recommended by 17·5%: Africa, 30·1%; Europe, 20·5%; and Western Pacific, 15·3% (*P* <·01) (Table 3).

#### Shoe cover

More than half of respondents (58·9%) reported that guidelines recommended the use of some type of shoe covers. Of note, this was higher in Southeast Asia (70·3%) than in other regions (*P* <·01) (Table 3).

### Training

Training sessions for HCWs concerning the appropriate and safe use of PPE were reported to happen by most respondents (235; 86·2%; 95% CI, 82·6-90·7) (Table 2, Fig. 1) with higher frequencies reported in HICs (74/79; 93·6%; 95% CI, 88·3-99) and UMICs (83/88; 99%; 95% CI, 89·4-99·1) than in LICs (30/48; 62·5%; 95% CI, 62·5-76·2) (*P* <·01) (figure 2).

**Figure 2:**
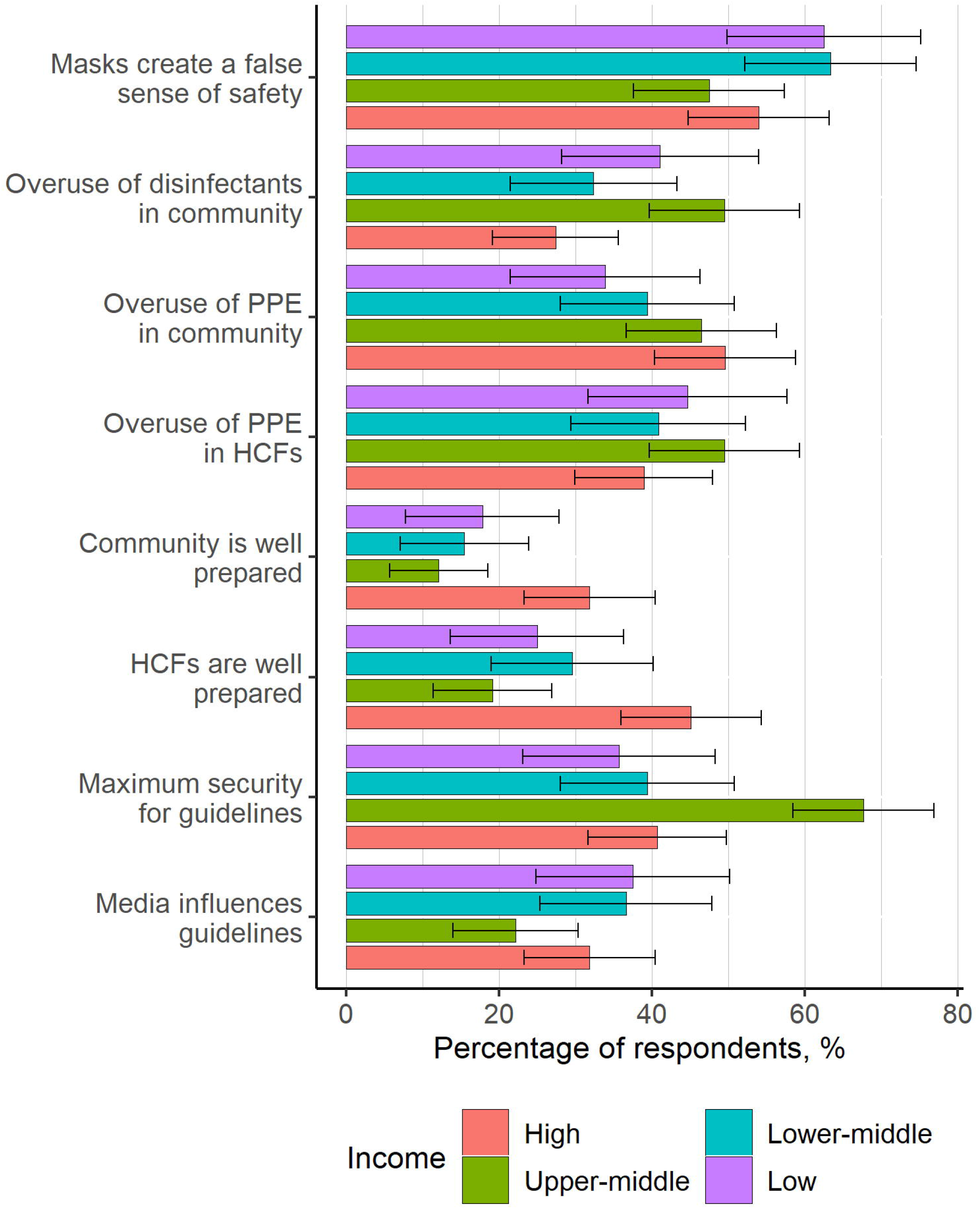
Perceived Opinion of the Level of Preparedness of Institutions for COVID-19 Among Survey Respondents, Stratified by Country Income Level. HIC, high-income country; UMIC, upper-middle income country; LMIC, lower-middle income country; LIC, low-income country; PPE, personal protective equipment; IPC, infection prevention and control.

### COVID-19 and PPE supplies

When asked about the availability of PPE supplies, 163 of 339 (48%; 95% CI, 42·7-53·4) respondents reported a shortage of supplies (64·2% [36/56; 95% CI, 51·7-76·8] in LICs compared with 27·4% [31/113; 95% CI, 19·2-35·6] in HICs). Shortages were reported commonly in Southeast Asia (51/72; 70%; 95% CI, 60·3-81·3) and Africa (65/113; 57·5%; 95% CI, 48·4-66·6). Shortage of facemasks was reported across all regions (Africa, 25·6%; the Americas, 21·2%; Eastern Mediterranean, 6·6%; Europe, 20·6% and Southeast Asia, 37·5%), except the Western Pacific (0%). Additional PPE supplies have been ordered across all regions (Africa, 19·4%; the Americas, 30·3%; Eastern Mediterranean, 40%; Europe, 33·7%; Southeast Asia 73·6%; and the Western Pacific 64·2%). However, increasing the PPE stockpile as part of the COVID-19 preparedness plan was only reported in 12% of institutions (41/339), with more in HICs (19%; 22/113) than LICs (7%; 4/56). Distribution of PPE supplies was organized by the government (i.e. ministry of health) in 24·1% of institutions (82/339), while only 19·4% (66/339) of facilities could order as much as necessary. Of note, 44·2% (150/339) were not aware of the distribution procedure of PPE supplies at their institution.

### Environmental Decontamination

Most respondents (199/270; 73·7%; 95% CI, 68·4-78·9) included hypochlorite disinfectant for environmental decontamination. Approximately one-third (100/270; 37·4%; 95% CI, 31·2-42·8) used automated room disinfection system technology at their institutions, such as hydrogen peroxide vapor and ultraviolet light (Table 2). Among these, 12·5% (6/48; 95% CI, 3·1-21·8) were in LICs compared with 35·9% (28/78; 95% CI, 25·2-46·5) in HICs (figure 1).

### Community Preparedness

Less than one-third (69/253; 27·2%; 95% CI, 21·7-32·7) believed that community preparedness was adequate, with the lowest frequencies reported in Southeast Asia (9/63; 14·2%) and Africa (18/87; 20·6%) (*P* = ·003), and a higher preparedness in HIC (36/71; 50·7%) than in LICs (10/45; 22·2%) (*P* <·01). More than half (144/270; 53·3; 95% CI, 47·3-59·2) of respondents reported that PPE use for COVID-19 was recommended in the community, mainly in Southeast Asia 77·2% (figure 2).

### Overuse/misuse of supplies

More than half (149/253; 58·8%; 95% CI, 52·3-64·9) of respondents agreed that there was an overuse/misuse of PPE, with a significant variation according to region (*P* < ·05) and income level (*P* < ·001). Among these, 74·3% (188/253; 95% CI, 68·9-79·6) believed that the use and heightened focus on wearing facemasks creates a misplaced feeling of safety, possibly reducing attention on other IPC measures, such as hand hygiene. Fifty percent of respondents (95% CI 43·6-55·9) agreed that there was a possible overuse/misuse of disinfectants in the community (figure 2).

### Media exposure

The belief that opinions expressed by the media influenced the choices made for national/local guidelines or the preparedness plans for COVID-19 was confirmed by 41·5% (105/252; 95% CI, 35·4-47·5) of respondents. More than half (161/253; 63·6%; 95% CI, 57·7-69·5) also believed that national/local guidelines were based predominantly on maximum security, rather than on evidence-based recommendations. HICs were more likely than LICs to report sufficient preparedness (51/71; 71·8%; 95% CI, 61·3-82·2 vs 14/45; 31%; 95% CI, 17·5-44·6; *P* <·01) (Fig.2).

## Discussion

The COVID-19 pandemic has raised international concern prompting IPC professionals to better control transmission and mitigate potential consequences. Our exploratory survey was initiated less than 4 weeks after WHO’s Public Health Emergency of International Concern declaration on 30 January 2020 to provide insight into the state of COVID-19 IPC preparedness worldwide. The vast majority of respondents believed that preparedness guidelines were based on safety-maximized procedures rather than on evidence-based recommendations; thus, uncertainties regarding the transmission modes of COVID-19 continue to generate controversy.[1],[15]

Our results show a wide variation of PPE recommendations, with predominantly the use of N95/FFP2 respirators only or of surgical masks only or in combination with respirators in specific situations. At the height of the outbreak, uncertainties about transmission led many institutions to impose airborne precautions while considerable variation was observed amongst international guidelines. The main transmission modes of COVID-19 virus occur via respiratory droplets and contact [2,11,16] and contact and droplet precautions were effective to control the MERS-CoV outbreak in a haemodialysis unit in Saudi Arabia[10] and SARS in Hong Kong.[17],[18] Aerosol-generating procedures have been associated with the incidence of transmission of coronaviruses.[19] Mainly guidelines do not recommend the use of shoe covers and the transmission risk from contaminated footwear is likely low.[20] However, PPE guidelines for HCWs in China recommend the use of shoe covers.[21] More uniformity is needed at the international level on PPE recommended for care of suspected or confirmed COVID-19 patients, based on available evidence and the most effective IPC strategies.

At the time of the survey, most respondents were engaged in providing training to HCWs on the appropriate and safe use of PPE, although lower frequency of training were reported in LMICs. Recent healthcare-associated infections and deaths among HCWs in China[12] and Italy[13] have emphasized the importance of training in appropriate PPE use, particularly in performing a fit test of a particulate respirator, safe PPE doffing and appropriate hand hygiene and use of gloves.[22] SARS infection transmission among HCWs was associated with an inadequate compliance with PPE, lack of understanding of IPC procedures and less than 2 h of infection control training.[8] Although the importance of fit testing has been highlighted, national standards vary widely and few documents explain the procedure in detail.[23] In some countries, fit testing is required anually or before use, while in others it is not a requirement, or fully absent, even though it has been shown that the efficacy of respirator use improves after fit testing is performed [24].

Hands-on training sessions on the use and compliance with PPE precautions and hand hygiene compliance monitoring have been recommended as key measures.[22] HCWs not trained in safe PPE doffing run the risk of accidental self-contamination, as reported during the Ebola outbreak.[9] Innovative approaches to training are important to mitigate the risk of viral self-contamination, suggesting that a rigorous process must be designed, combined with standardized training.[25,26] The PPE doffing process is critical to keep HCWs safe and further research on the science of human factors and HCW behavior with respiratory protection safety is needed.

Shortage of PPE has become a global concern; 48% of respondents reported a shortage of PPE across all regions, notably facemasks, with higher frequencies in LMICs. Importantly, this shortage jeopardizes outbreak response strategies by impeding appropriate and safe action to combat COVID-19 spread. An increasing need for PPE supplies worldwide remains unmet and places HCWs and patient safety under threat. Protection of HCWs should be prioritized and healthcare facilities must have the necessary equipment. Political leaders and health authorities need to urgently seek solutions and develop contingency plans to reserve and allocate resources where these are needed the most. Increasing the PPE stockpile as part of the COVID-19 preparedness plan was only reported in 12% of facilities. While HICs have established IPC programmes and yet are faced with deficient supplies, LICs may not have the resources necessary to fight the current pandemic. WHO is undertaking major efforts to coordinate the Emergency Global Supply Chain System facilitating the purchase and shipment of PPE and medical supplies across the globe; as of 7 April, millions of pieces of PPE were shipped to 133 countries.[27] Local production of alcohol-based hand rub using the WHO-recommended formulation[28] has been reported in many countries in an attempt to replace the current shortage. Some facilities are also considering reprocessing and reuse of single-use PPE.[29] WHO has provided guidance of rational use of PPE including considerations pertaining to reprocessing and reuse in case of severe shortage.[30]

The survival of coronaviruses on environmental surfaces and the impact of environmental contamination on healthcare-associated outbreaks as a mechanism of transmission argue for enhanced environmental hygiene.[20,31,32] A chlorine-based surface disinfectant was used by most participating facilities, but there appears to be an infrequent use of automated room disinfection system technology. Automated disinfection system technology may be more accurate as it diminishes reliance on the operator.[33] Decontamination of high-touch surfaces as a potential source of transmission needs to be considered.

## Limitations

In the context of global health with increased attention to the COVID-19 epidemic evolving and a situation that is rapidly changing, our specific intent was to provide a snapshot of the state of affairs of IPC implemented measures less than four weeks after WHO’s declaration of a public health emergency of international concern and while it had been declared a global pandemic. Since then, IPC preparedness measures in healthcare facilities across countries have intensified and therefore it will be interesting to assess with a follow-up survey.

Our study has limitations. First, this is a voluntary survey consisting of self-selected participants whose perceptions and opinions might not be entirely representative for healthcare facilities of a country and there was no validation method of the participants’ data entry. Second, most responses were from tertiary-care healthcare facilities, which might suggest an overrepresentation of these settings. However, tertiary-care hospitals have a central role in preparedness activities for emerging infectious diseases and our results provide important information on their state of preparedness. Third, some of the questions in our survey remained unanswered (‘I don’t know’) by a proportion of participants as depicted in Table 2 and 3, likely due to some topics not having relevance to the local context of study participants. Fourth, due to time and resource constraints the survey was made available only in English and Chinese. Finally, the questions of the survey were not formally validated, since the survey items were based mainly on the WHO COVID-19 global research roadmap priorities that had been identified. Regardless of these limitations, the study provides some important insight on necessary IPC measures that need to be considered to scale-up preparedness to combat COVID-19.

## Conclusion

Our findings, including identified concerns, may help to provide direction to improve preparedness in countries and prioritization of critical actions. The COVID-19 global pandemic has shown the importance of building more resilient healthcare systems with effective IPC as key to avoid or mitigate outbreaks impact. Results revealed lack of guidelines and, when available, large variations exist in recommendations. Health organizations should jointly evaluate the available evidence and develop a uniform policy on the appropriate PPE to be used. Strengthening of coordinated international efforts is urgent to address the challenges related to the major PPE shortage in healthcare facilities, particularly the lack of resources in low-income settings, and to improve reliable communication through the media.

## Data Availability

All relevant data are within the manuscript. Any other information needed is available upon request.

## Acknowledgments

We are greatly indebted to all colleagues and members of the International Society of Antimicrobial Chemotherapy on Infection Prevention and Control (ISAC-IPC) Working Group and Infection Control Africa Network (ICAN) who participated in this survey and for contributions to the questionnaire.

## Contributors

ET, JH, BA and AV were involved in study design and implementation. ET and JH contributed equally to the work. ET, JH, BG, KM, VCCC, SCW, AW, FO, AV undertook data collection. ET and JH did data analysis and made the figures and tables. ET, JH, BA, BG, AW, VCCC, SCW, KM, FO and AV were involved in data interpretation and study coordination and contributed to writing and reviewing the manuscript. All authors approved the final version for publication.

## Disclaimer

The opinions expressed in this Article are those of the authors and do not reflect the official position of WHO. WHO takes no responsibility for the information provided or the views expressed in this article.

## Ethical approval

This study was exempted by the Radboud University Medical Center (The Netherlands) as it did not fall within the remit of the Medical Research Involving Human Subjects Act (NL2020-6262).

## Supplementary File

**1. STROBE checklist**.

**2. Survey Instrument**.

